# COVID-19 Risk Perception Among U.S. Adults: Changes from February to May 2020

**DOI:** 10.1101/2020.08.20.20178822

**Authors:** Amyn A. Malik, SarahAnn M. McFadden, Jad A. Elharake, Obianuju Genevieve Aguolu, Mehr Shafiq, Saad B. Omer

## Abstract

The COVID-19 pandemic continues to detrimentally impact the United States. Using a survey, we collected demographic and COVID-19 risk perception, behavior, knowledge, and attitude data from 672 adults across the U.S. in May 2020. These variables were compared with the results from a survey in February 2020. Participants who were older (55+ years; M = 6.3, SD = 2.0), identified as Native American/Alaska Native (M = 6.8, SD = 1.0) or Asian (M = 6.0, SD = 2.0), and those who had contracted (M = 6.8, SD = 2.0) or knew someone who had contracted COVID-19 (M = 6.2, SD = 1.7) reported higher perceived risk. Health behaviors, such as physical distancing, have shown to impact infectious disease trajectories. As the U.S. reopens its economy, public health officials and politicians must formulate culturally appropriate and evidence-based messaging and policies, based on the public’s COVID-19 risk perceptions, to encourage preventive behaviors.

## Introduction

In less than six months, the coronavirus disease 2019 (COVID-19) pandemic has become the most challenging global health pandemic since the 1918 flu.^1^ Despite local and state governments implementing measures to reduce the spread of the virus, the COVID-19 pandemic continues to threaten human life and the immediate return to normalcy. As of August 20, 2020, the U.S. leads the world in confirmed cases (5.5 million; 1.7% of U.S. population) as well as total number of confirmed deaths (175,000).^2^ Although many Americans are following the Centers for Disease Control and Prevention’s (CDC) COVID-19 guidelines, such as physical distancing, wearing a mask, and frequent hand washing, others have decided to forgo these safe and protective measures.^3^ Americans typically make health decisions by weighing risk for consequences with benefits of action, which necessitates understanding risk perception trends across the country.^4^ This need is further elevated because the number of COVID-19 cases and deaths continues to rise in the U.S. Understanding the societal and cultural factors that drive public health risk perception and associated health outcomes, therefore, becomes a high priority.

Studies conducted on previous pandemics demonstrate strong evidence on how compliance with health behaviors recommended by public health officials can significantly influence the trajectory of an outbreak.^5–13^ With COVID-19 vaccines currently under development and not likely to be available for at least several months,^14^ U.S. public health officials and politicians must evaluate the public’s risk perceptions of COVID-19 to formulate strategic messaging and policies in hopes of increasing safe health behaviors and effectively controlling the COVID-19 pandemic. Additionally, while all Americans are impacted by COVID-19, vulnerable populations are facing a disproportionate burden of cases and deaths.^15^ Due to social and structural determinants of health and inequitable access to healthcare,^16^ current data show the most alarming disparities are observed among low-socioeconomic populations and African American, Hispanic, and Native American communities.^17–22^ While considering this nuance, public health and political leaders must also develop culturally competent messaging strategies, especially given the medical community’s history with conducting unethical studies within underrepresented communities,^23^ that cater to vulnerable populations to mitigate these outcomes and promote health equity.

As the COVID-19 outbreak continues to evolve and the U.S. begins to gradually reopen its economy, the purpose of our study is to understand the social and cultural factors that influence COVID-19 risk perception among the adult U.S. population and how it has changed from the early months of the pandemic. This will provide the public health community with the necessary information needed to better inform messaging interventions.

## Results

### Demographic Differences in Risk Perception

The sample consisted of 672 out of 938 eligible adults (completion rate: 72%). The sample was fairly representative of the U.S. population in terms of age, gender, race, ethnicity, and education (Table 1). The majority of participants were non-Hispanic white (n = 436, 65%) and had a college or graduate degree (n = 351, 52%). The average risk perception score was 5.9 (SD = 2.0). When compared to results from our February 2020 survey, participants from the May survey perceived they were at higher risk (MD = 0.91, p < 0.01) of contracting the coronavirus. However, risk perception varied based on several different factors (Table 1). Older adults had a high risk perception score (M = 6.3, SD = 2.0; Table 1) Additionally, participants who reported being sick COVID-19 had a high risk perception score (M = 6.8, SD = 2.0; Table 1).

**Table 1:** Participants’ Risk Perception During the COVID-19 Pandemic (May 2020)

|  | Total<br>(N = 672) n<br>(%) | Risk Perception Score<br>(out of 10;<br>M +/- SD) | U.S. Population<br>(%) |
| --- | --- | --- | --- |
| Age (years) |  |  |  |
| 18-25 | 73 (11) | 5.4 +/- 2.2 | 12 |
| 26-35 | 107 (16) | 5.5 +/- 2.0 | 18 |
| 36-45 | 141 (21) | 5.9 +/- 2.0 | 16 |
| 46-55 | 95 (14) | 5.4 +/- 2.0 | 17 |
| 55+ | 256 (38) | 6.3 +/- 2.0 | 36 |
| Race |  |  |  |
| Black/<br>African American | 67 (10) | 5.4 +/- 2.1 | 13 |
| Native American/Alaska<br>Native | 19 (2.8) | 6.8 +/- 1.0 | 1.0 |
| Asian | 97 (14) | 6.0 +/- 2.0 | 5.0 |
| Native Hawaiian/<br>Other Pacific<br>Islander | 2 (0.20) | 5.5 +/- 1.0 | 0.0 |
| White | 487 (73) | 5.9 +/- 2.0 | 73 |
| Ethnicity |  |  |  |
| Hispanic | 68 (10) | 6.2 +/- 1.7 | 18 |
| Non-Hispanic | 604 (90) | 5.8 +/- 2.0 | 82 |
| Education |  |  |  |
| No High School | 10 (2) | 6.1 +/- 2.5 | 12 |
| High School | 162 (24) | 5.5 +/- 2.0 | 27 |
| Some College | 149 (22) | 6.0 +/- 2.0 | 29 |
| College | 205 (30) | 6.0 +/- 2.0 | 19 |
| Graduate/Professional | 146 (22) | 6.0 +/- 2.0 | 12 |
| Employment Status |  |  |  |
| Employed | 314 (47) | 5.8 +/- 2.0 | 51 |
| Unemployed | 194 (29) | 5.7 +/- 2.0 | 15 |
| Due to COVID-19 | 57 (29) | 5.6 +/- 1.8 | * |
| Not Due to COVID-19 | 137 (71) | 5.7 +/- 2.2 | * |
| Retired | 164 (24) | 6.3 +/- 1.8 | * |
| Sick with COVID-19 |  |  |  |
| Yes | 20 (3.0) | 6.8 +/- 2.0 | 0.7 |
| No | 652 (97) | 5.8 +/- 2.0 | 99.3<sup>42</sup> |
| Someone you know who is sick with COVID-19 |  |  |  |
| Yes | 138 (21) | 6.2 +/- 1.7 | * |
| No | 534 (79) | 5.8 +/- 2.1 | * |
| Chronic Illness |  |  |  |
| Yes | 162 (24) | 6.6 +/- 1.8 | 60<sup>43</sup> |
| No | 510 (76) | 5.7 +/- 2.0 | 40 |

\*unable to find U.S. adult population estimates for these categories

A multivariable linear regression analysis supported these descriptive findings. When controlling for other variables, adults over 55 years of age reported higher risk perception than younger adults (β = 0.9, p = 0.01; S Table 1). Participants who reported being Native American/Alaska Native (β = 1.54, p = 0.03) and Asian (β = 0.87, p = 0.02) also reported increased risk perception compared to participants who reported being Black/African American (S Table 1). Finally, participants who knew someone who had been sick with COVID-19 in the past or at the time the survey was conducted had an increased risk perception score compared to those who did not. (β = 0.55, p = 0.01; S Table 1).

### Knowledge

Approximately 93% (n = 627) of our sample reported that they were aware of the COVID-19 pandemic. When asked to rate their own knowledge about COVID-19, 72% (n = 481) of our sample said they had good or very good knowledge about COVID-19 compared to 4% (n = 24) who said they had poor or very poor knowledge about COVID-19. When we conducted this survey in February 2020, 39% (n = 281) of our sample said they had good or very good knowledge about COVID-19, and 17% (n = 119) said they had poor or very poor knowledge about COVID-19 (p < 0.01).

When asked about whether there was a cure or a vaccine for COVID-19, 81% of our sample (n = 541) correctly answered that there currently was not. However, of the 13% who answered incorrectly, 9% (n = 61) thought there was a curative treatment, and 4% (n = 26) thought there was a vaccine. When asked where they learned about the curative treatment or vaccine, around 50% of participants said they got their information from the media on television (treatment: 48%, n = 29; vaccine: 54%, n = 14).

A knowledge score was calculated based on 18-items. On average, the participants from the second survey administered in May scored higher on the knowledge questions compared to the participants from the first survey administered in February (MD = 0.85, p < 0.01; Figure 1).

**Figure 1.**
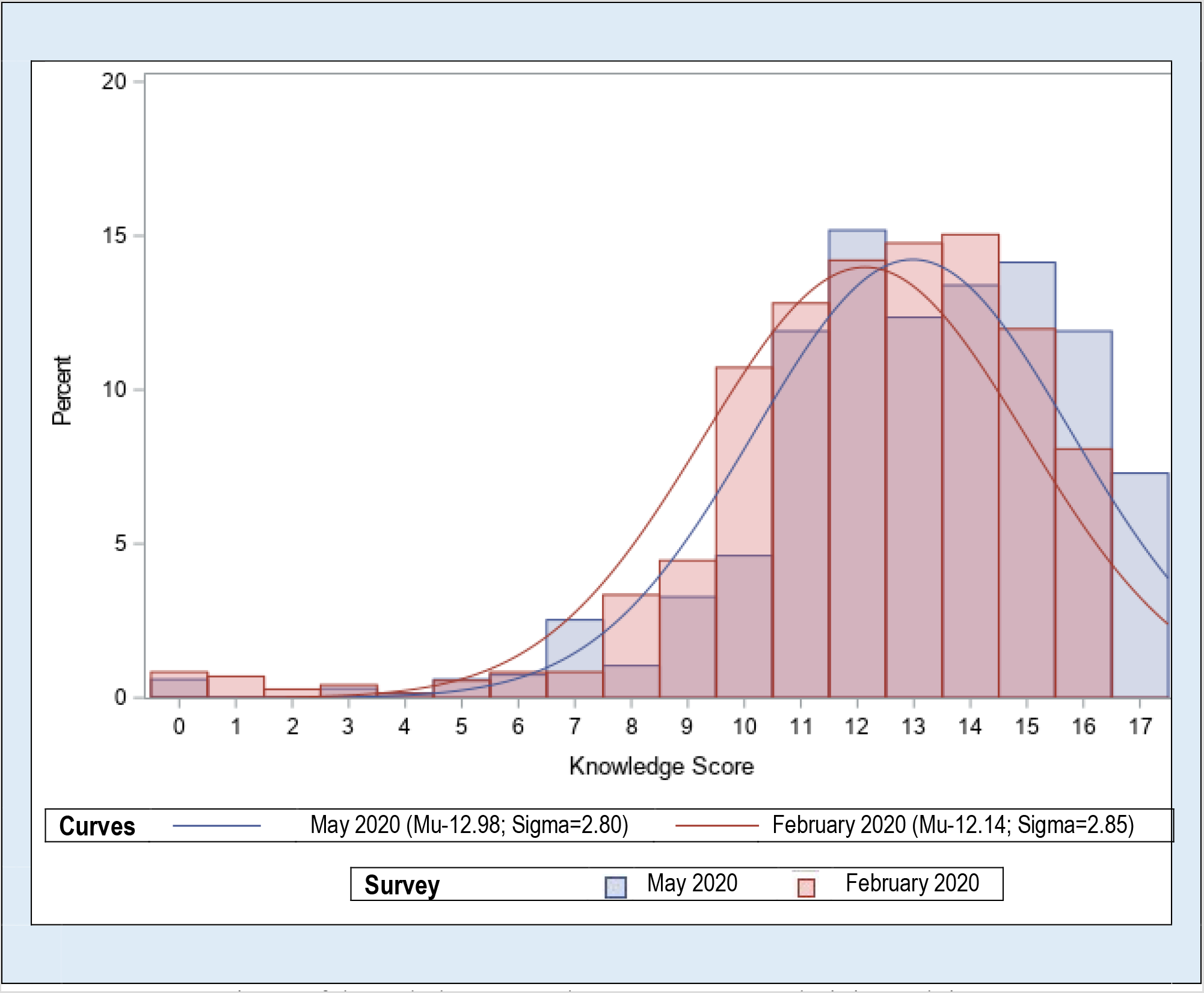
Comparison of knowledge score between survey administered in May to survey administered in February

### Infection Prevention Policy and Practices

Strict policies for infection prevention including quarantine, travel restrictions, closing community facilities, and cancelling major events were endorsed by most participants (Table 2). Participants that agreed or strongly agreed with these policies had a higher risk score than participants who disagreed with these policies (Table 2). Interestingly, 36% of participants agreed or strongly agreed that non-violent inmates should be released from jail or prison to decrease crowding and slow the pandemic (Table 2). Those who agreed with releasing inmates had a higher risk perception score (M = 6.4, SD = 1.8) than those who disagreed (M = 5.6, SD = 2.1). Additionally, risk perception was higher (M = 6.2, SD = 1.9) in the 30% of participants who agreed that in the case of a pandemic it is reasonable to “temporarily discriminate” against a community based on their country of origin than in the 70% (M = 5.7, SD = 2.1) who did not agree with this statement.

**Table 2:**
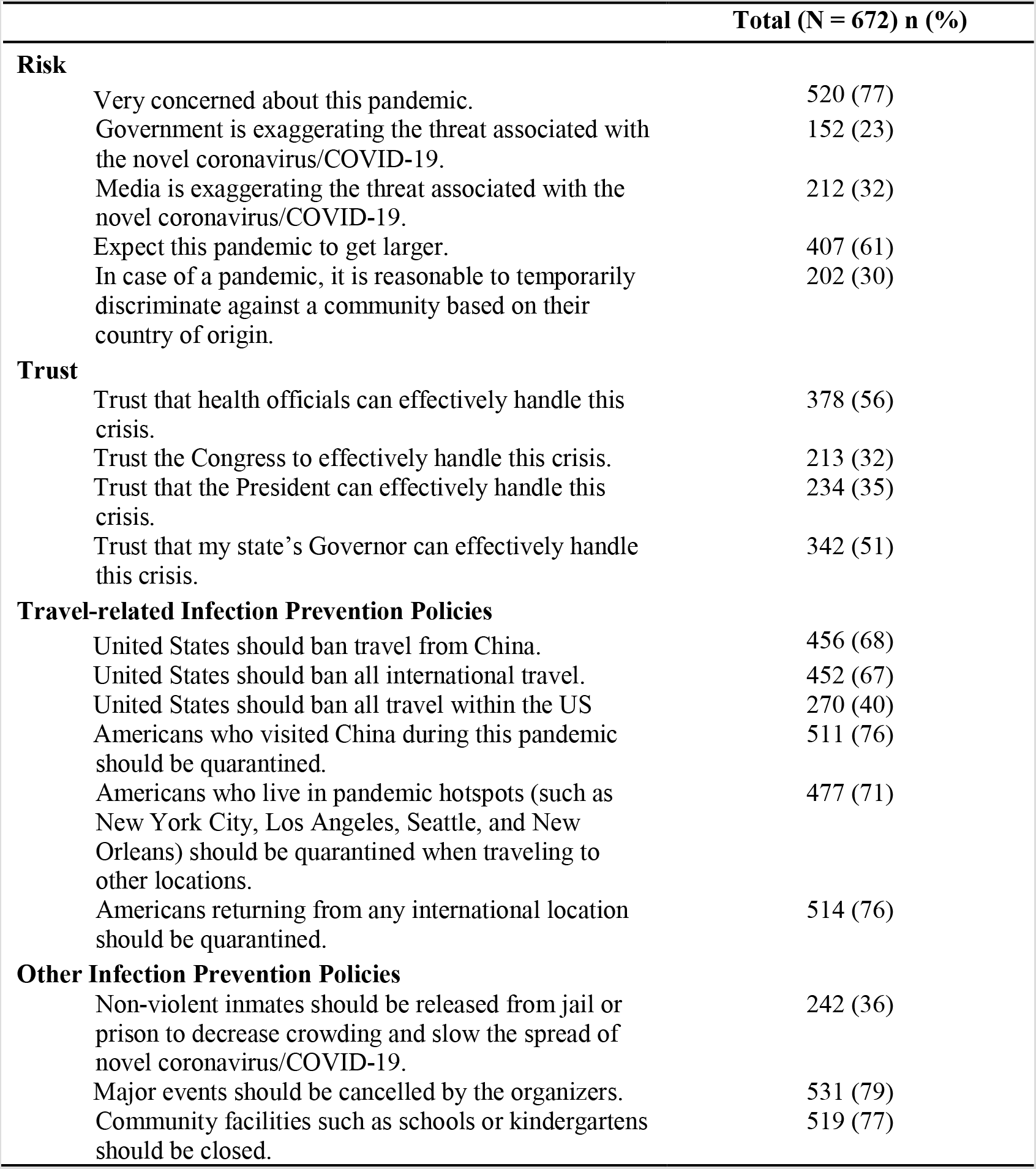
Participants’ Attitudes regarding COVID-19 Risk, Trust, Travel, and Infection Prevention Polices in May 2020.

|  | Total (N = 672) n (%) |
| --- | --- |
| **Risk** |  |
| Very concerned about this pandemic. | 520 (77) |
| Government is exaggerating the threat associated with the novel coronavirus/COVID-19. | 152 (23) |
| Media is exaggerating the threat associated with the novel coronavirus/COVID-19. | 212 (32) |
| Expect this pandemic to get larger. | 407 (61) |
| In case of a pandemic, it is reasonable to temporarily discriminate against a community based on their country of origin. | 202 (30) |
| **Trust** |  |
| Trust that health officials can effectively handle this crisis. | 378 (56) |
| Trust the Congress to effectively handle this crisis. | 213 (32) |
| Trust that the President can effectively handle this crisis. | 234 (35) |
| Trust that my state's Governor can effectively handle this crisis. | 342 (51) |
| **Travel-related Infection Prevention Policies** |  |
| United States should ban travel from China. | 456 (68) |
| United States should ban all international travel. | 452 (67) |
| United States should ban all travel within the US | 270 (40) |
| Americans who visited China during this pandemic should be quarantined. | 511 (76) |
| Americans who live in pandemic hotspots (such as New York City, Los Angeles, Seattle, and New Orleans) should be quarantined when traveling to other locations. | 477 (71) |
| Americans returning from any international location should be quarantined. | 514 (76) |
| **Other Infection Prevention Policies** |  |
| Non-violent inmates should be released from jail or prison to decrease crowding and slow the spread of novel coronavirus/COVID-19. | 242 (36) |
| Major events should be cancelled by the organizers. | 531 (79) |
| Community facilities such as schools or kindergartens should be closed. | 519 (77) |

Most participants reported it was easy to follow the CDC public health recommendations (n = 395, 59%). For the 115 participants (17%) who reported it was hard to follow the CDC guidelines, the most common reasons were “We don’t know how long this pandemic is going to last” and “I’m classified as an essential worker” (Table 3). Excluding the use of facemasks, over 90% of our sample reported following CDC guidelines for infection prevention (S Figure 1). For facemasks specifically, 85% (n = 571) of our sample reported using them.

**Table 3:** Participants Reporting Difficulty in following in Following CDC/Public Health Recommendations.

| Why is it hard to follow the CDC/public health recommendations? | No. | (%) |
| --- | --- | --- |
| We don't know how long this pandemic is going to last. | 32 | (28) |
| I'm classified as an essential worker. | 25 | (22) |
| I have bills that need to be paid. | 18 | (16) |
| The recommendations are not clear. | 11 | (10) |
| Other | 10 | (9) |
| I can't get basic supplies needed for cleaning my home. | 7 | (6) |
| My family relies on me as a caretaker. | 7 | (6) |
| I live in a small apartment. | 5 | (4) |

Categories are mutually exclusive
Participants who reported a difficulty in following these recommendations (n = 115, 17.1%) are a part of a subset of the overall sample (n = 672)

Participants who practiced or planned on buying extra medicines, food and other everyday items, and disinfectants had a statistically significant higher risk perception as compared to those who did not (S Table 2).

### Public Health and the Economy

Regardless of employment status, most participants agreed that the COVID-19 pandemic has severely damaged the economy (n = 566, 84%) and that slowing the spread of COVID-19 is more important than the economy (n = 457, 68%). Additionally, 70% (n = 473) of participants said that public health should be prioritized over opening the economy (S Table 3).

### Confidence in Sources of Information and Leadership

Overall confidence in healthcare professionals (M = 4.2, SD = 1.1) and health officials (M = 4.2, SD = 1.2) was high, and confidence in social media was low (M = 2.9, SD = 1.5; S Figure 2). With respect to the COVID-19 pandemic specifically, participants reported high confidence in information from health professionals (M = 4.2, SD = 1.0) and their own physician (M = 4.2, SD = 1.1; S Figure 2). Participants reported the lowest confidence in COVID-19 pandemic information from the White House (M = 2.9, SD = 1.6) and Congress (M = 2.8, SD = 1.4; S Figure 2). When compared to the survey administered in February 2020, there was significant reduction in the reported confidence levels for CDC, DHHS, NIH, and other professional organizations such as AMA (S Table 4).

We asked participants to rank who they wanted the U.S. COVID-19 response to be led by, and 35% (n = 237) of participants ranked the president as their first choice to lead the pandemic response (S Figure 3). This is in contrast the survey results in February 2020 where 13% (n = 91) of participants wanted the president to lead the response, (p < 0.01). Comparatively, 17% (n = 117) of participants wanted the CDC to lead the response, in contrast to the 53% (n = 382) who wanted the CDC to lead the response when surveyed in February (p < 0.01; S Figure 3).

As sensitivity analysis, we carried out a weighted analysis for COVID-19 risk perception using U.S. demographics. Data for weighted analysis were extracted from U.S. Census data.^24^ Weighting by age, gender, and race decreased the mean risk perception score from 5.9 to 5.2 and increased the percentage of population that supports “temporary discrimination” based on nationality from 30 to 38.

## Discussion

While COVID-19 risk perceptions are high among the U.S. adult population, there is a great deal of variability among different demographic groups. Additionally, compared to our previous survey conducted in February 2020, a greater proportion of participants reported having an above average perceived knowledge about COVID-19. Although currently there are treatments, such as dexamethasone and remdesivir, that improve outcomes in patients experiencing COVID-19, at the time the survey was conducted there were no scientifically accepted treatments. However, over 15% of our participants thought there was a curative treatment or vaccine for COVID-19 and learned this information from television media. This finding is concerning and likely due to the increased level of misinformation and inconsistent messaging among various news outlets.^25^ Therefore, our study demonstrates the continued need for targeted messaging about appropriate infection prevention and protective measures. Understanding COVID-19 risk perception and knowledge may help public health agencies anticipate and address potential barriers to preventive behavior to avert thousands of more infections and deaths by COVID-19.^26^

There is strong evidence that a perceived lack of consistent information during a crisis may lead to distrust, lack of confidence, and fear among the public.^27^ While 35% of our participants ranked the president as their first choice to lead the pandemic response, participants reported the lowest confidence in COVID-19 pandemic information from the White House and Congress. Confidence in healthcare professionals and health officials was the highest; however, there was a significant reduction in the reported confidence level in the CDC compared to the February 2020 survey. These findings may be due to the contradicting COVID-19 information and lack of immediate response to the crisis from the White House, Congress, and CDC.^28^ The White House, Congress, and public health organizations, especially the CDC, must agree on the information being communicated to the public and be transparent with the American people to ensure trust in public health and governmental authorities.

There is great responsibility on politicians and healthcare leaders to create messaging that is clear and consistent, as well as evidenced-based and culturally competent. However, currently, there is evidence that non-Asian Americans’ fear visiting local communities with higher concentration of Asian Americans, which is reflected in their hesitance in ordering from Asian-owned eateries.^29^ This may be a result of fear in a subgroup of the population that there are greater chances of contracting COVID-19 from Asian-Americans than non-Asian Americans. Moreover, this notion is fueled by political leaders who repeatedly claim that China is responsible for the pandemic.^30^ Our finding that 30% of our participants agreed that it is reasonable to “temporarily discriminate” against a community based on their country of origin in the case of a pandemic supports this; those who agreed with “temporary discrimination” reported a higher risk perception than those who disagreed.

Irrespective of employment status, 68% of our participants agreed that slowing the spread of COVID-19 was more important than the economic disruption. Furthermore, most of our sample supported the closure of community facilities and schools as well as the cancellation of large social events. This contrasts with the common narrative depicted by the media that most Americans are against the lockdown, and in doing so, the media may be spreading a narrative not supported by the majority.^31^ Messaging strategies must build awareness and knowledge of risks and benefits of prevention methods, especially considering that 17% of our participants reported having difficulties following the CDC COVID-19 guidelines. These participants mostly consisted of workers classified as essential or those unable to pay their bills, which also reflects the detrimental economic impact of COVID-19. Public health leaders and politicians must identify areas of citizens’ concerns and alleviate them through targeted messaging, and strongly consider providing more financial support for Americans.

Our findings may be influenced by possible selection bias because participants needed a CloudResearch account and access to a smartphone or computer to participate. We had a response rate of 72% and our data are fairly representative of the U.S. adult population. Another strength of our study includes timeliness. This is also one of the first studies that looks at detailed COVID-19 risk perceptions and associated health behaviors. Moreover, due to it being a longitudinal study, we were able to identify changes in risk perception, behavior, attitude, and messaging needs among the U.S. adult population since the beginning of the COVID-19 pandemic.

Consistent with previous literature, our study calls on the importance of building trust and understanding the risk perception landscape among the U.S. public. It also highlights the need for evidence-based messaging of recommended prevention measures to mitigate crises during the COVID-19 pandemic.

## Methods

### Participants and procedure

Data were collected using an electronic questionnaire via Qualtrics® (Qualtrics, Provo, UT). In early May 2020, participants completed a questionnaire on CloudResearch.^32^ CloudResearch is an online survey platform that allows for representative surveying. The goal of our sampling was to be representative of the U.S. general population based on age, gender, education, race and ethnicity. Participants were eligible if they were 18 years of age or older, could read English, and had a CloudResearch account with access to the internet via computer or smartphone. Participants received compensation in the amount they agreed to with the platform through which they entered this survey.

### Measures

Basic demographic information was collected as well as zip code, state of residence, and employment status. Participants also completed the perceived risk scale (Cronbach’s α = 0.72) which had 10 survey-items (5-point Likert Scale: 0 = strongly disagree/disagree/neutral; 1 = agree/strongly agree). Variables used for risk perception score calculation are shown in S Figure 5). Participants were removed from score calculation if they selected “don’t know” to one or more of the items on the risk perception scale. For scoring, variables collected on Likert Scale were dichotomized as: 0 = strongly disagree/disagree/neutral; 1 = agree/strongly agree. Additionally, participants were asked about their knowledge of COVID-19, effective preventive measures, measures being practiced, confidence and the reliability of sources of information regarding the COVID-19 pandemic (5-point Likert Scale: 1 = strongly disagree to 5 strongly agree), support for restrictive infection prevention policies, and impact of COVID-19 on their employment and behavior. Finally, participants were asked to rank who they thought should lead the U.S. response to the COVID-19 pandemic. The full survey instrument is included as a supplement. This survey instrument was adapted from McFadden et al^14^ and COSMO standard protocol for WHO.^33^

A knowledge score consisting of 17 survey-items was constructed by adding all the points for correct items for each participant (Cronbach’s α = 0.78; S Table 6). For scoring, variables collected on Likert Scale were dichotomized as: 0 = strongly disagree/disagree/neutral; 1 = agree/strongly agree.

To compare the current risk perception and calculate change over time, we used data collected from a similar survey in early February 2020. Details of this study are reported here.^34^

### Statistical Analysis

Descriptive statistics (frequencies, percentages) were calculated for the sample demographic characteristics, trust, attitude, and behavior of the participants. T-test and one-way ANOVA for continuous variables and chi-square test for categorical variables were used to compare the change in risk perception, knowledge, attitude, and behavior between the surveys in February and May 2020 using variables collected at both time points.

To test the associations between demographic factors and exposure to COVID-19 with COVID-19 risk perception, a multi-variable linear regression analysis was used.

Data were analyzed using Stata Version 16 (StataCorp, College Station, Texas).

### Ethical Approval

Yale University Institutional Review Board approved this study (IRB protocol number: 2000027891). Participants provided informed consent prior to data collection.

### Data Availability

The raw data will be accessible on contacting the corresponding author.

## Data Availability

Data is available by contacting corresponding author.

## Author’s contribution

SMM, AAM, and SBO conceptualized the study. SMM, and AAM collected data under supervision from SBO. SMM, AAM, JE, MS, and OGA performed and reviewed the analysis, and wrote the initial draft of the manuscript. All authors helped interpret the findings, read and approved the final version of the manuscript.

## Funding

The study was funded by the Yale Institute for Global Health.

## Declaration of Interests

All authors declare no conflict of interest.

## Extended Data

**S Table 1:** Predictors of risk perception related to COVID-19 in the US in May 2020 – Linear regression analysis

| Variables | Unadjusted Model |  |  | Adjusted Model |  |  |
| --- | --- | --- | --- | --- | --- | --- |
|  | Coef. | [95%<br>Conf. Interval] |  | Coef. | [95%<br>Conf. Interval] |  |
| Age |  |  |  |  |  |  |
| 18-25 |  | *Ref* |  |  | *Ref* |  |
| 26-35 | 0.09 | -0.57 | 0.76 | -0.01 | -0.69 | 0.66 |
| 36-45 | 0.60 | -0.04 | 1.23 | 0.47 | -0.18 | 1.11 |
| 46-55 | 0.05 | -0.64 | 0.74 | -0.10 | -0.82 | 0.62 |
| 55+ | **0.88** | **0.30** | **1.47** | **0.70** | **0.03** | **1.37** |
| Race |  |  |  |  |  |  |
| Black/African American |  | *Ref* |  |  | *Ref* |  |
| Native American/Alaska Native | **1.41** | **0.04** | **2.78** | **1.38** | **0.03** | **2.72** |
| Asian | 0.65 | -0.07 | 1.37 | **0.93** | **0.22** | **1.63** |
| Native Hawaiian/Other Pacific Islander | 0.11 | -2.74 | 2.96 | 0.56 | -2.28 | 3.41 |
| White | 0.49 | -0.11 | 1.09 | 0.52 | -0.07 | 1.11 |
| Ethnicity |  |  |  |  |  |  |
| Hispanic |  | *Ref* |  |  | *Ref* |  |
| Non-Hispanic | -0.33 | -0.89 | 0.24 | -0.44 | -1.02 | 0.13 |
| Education |  |  |  |  |  |  |
| No High School |  | *Ref* |  |  |  |  |
| High School | -0.65 | -2.09 | 0.79 |  |  |  |
| Some College | -0.15 | -1.59 | 1.29 |  |  |  |
| College | -0.12 | -1.55 | 1.31 |  |  |  |
| Graduate/Professional | -0.13 | -1.57 | 1.32 |  |  |  |
| Employment |  |  |  |  |  |  |
| Employed |  | *Ref* |  |  | *Ref* |  |
| Unemployed | -0.12 | -0.52 | 0.29 | -0.13 | -0.55 | 0.28 |
| Retired | **0.52** | **0.10** | **0.95** | 0.12 | -0.40 | 0.64 |
| Sick with COVID-19 |  |  |  |  |  |  |
| No |  | *Ref* |  |  | *Ref* |  |
| Yes | 0.92 | -0.05 | 1.89 | 0.34 | -0.69 | 1.36 |
| Know someone sick with COVID-19 |  |  |  |  |  |  |
| No |  | *Ref* |  |  | *Ref* |  |
| Yes | **0.46** | **0.04** | **0.87** | **0.55** | **0.11** | **0.98** |
| Gender |  |  |  |  |  |  |
| Male |  | *Ref* |  |  | *Ref* |  |
| Female | 0.07 | -0.28 | 0.41 | 0.15 | -0.19 | 0.50 |
| Other | **-2.10** | **-4.09** | **-0.11** | **-1.98** | **-3.92** | **-0.04** |
| Chronic Illness |  |  |  |  |  |  |
| No |  | *Ref* |  |  |  |  |
| Yes | **0.94** | **0.55** | **1.33** | **0.75** | **0.34** | **1.17** |

NB: Bolded values were statistically significant at $p < 0.05$ ; Education excluded from adjusted model due to collinearity

**S Table 2:** Participants’ Risk Perception Score and Behavior During COVID-19 Pandemic (May 2020)

| We would like to know if you implemented<br>or plan to implement the following behaviors | Risk Perception Score (Out of 10) |  |  |  |
| --- | --- | --- | --- | --- |
|  | No. (%) | Mean | Std. Dev. | p-value |
| Bought extra medicines |  |  |  |  |
| • I already did that | 204 (37.99) | 6.25 | 1.88 |  |
| • I plan to do that | 65 (12.10) | 6.22 | 1.72 |  |
| • I don't plan to do that | 268 (49.91) | 5.51 | 2.12 |  |
|  | **537 (100.00)** | **5.87** | **2.01** | **<0.01** |
| Bought extra supplies at the pharmacy |  |  |  |  |
| • I already did that | 189 (35.20) | 6.24 | 1.87 |  |
| • I plan to do that | 72 (13.41) | 6.24 | 1.83 |  |
| • I don't plan to do that | 276 (51.40) | 5.53 | 2.10 |  |
|  | **537 (100.00)** | **5.87** | **2.01** | **<0.01** |
| Bought extra medications that I take regularly |  |  |  |  |
| • I already did that | 200 (37.24) | 6.24 | 1.89 |  |
| • I plan to do that | 66 (12.29) | 6.06 | 1.89 |  |
| • I don't plan to do that | 271 (50.47) | 5.56 | 2.10 |  |
|  | **537 (100.00)** | **5.87** | **2.01** | **<0.01** |
| Bought food supplies on a large scale |  |  |  |  |
| • I already did that | 258 (48.04) | 6.10 | 1.93 |  |
| • I plan to do that | 67 (12.48) | 5.97 | 1.91 |  |
| • I don't plan to do that | 212 (39.48) | 5.57 | 2.12 |  |
|  | **537 (100.00)** | **5.87** | **2.01** | **0.02** |
| Bought other everyday things on a large scale |  |  |  |  |
| • I already did that | 212 (39.48) | 6.07 | 1.87 |  |
| • I plan to do that | 88 (16.39) | 6.05 | 1.89 |  |
| • I don't plan to do that | 237 (44.13) | 5.63 | 2.16 |  |
|  | **537 (100.00)** | **5.87** | **2.01** | **0.05** |
| Bought disinfectants on a large scale |  |  |  |  |
| • I already did that | 185 (34.45) | 6.11 | 1.94 |  |
| • I plan to do that | 98 (18.25) | 6.05 | 1.76 |  |
| • I don't plan to do that | 254 (47.30) | 5.63 | 2.14 |  |
|  | **537 (100.00)** | **5.87** | **2.01** | **0.03** |

**S Table 3:** Participants’ Attitudes on the Impact of COVID-19 Pandemic on the Economy –Stratified by Employment Status

|  | **Total (N = 672) n (%)** |
| --- | --- |
| **Employed** | 314 (47) |
| Novel coronavirus/COVID-19 has severely damaged the economy | 269 (86) |
| Slowing the spread of the pandemic is more important than damage to the economy | 225 (72) |
| Public health should be prioritized over opening the economy | 229 (73) |
| **Unemployed** | 194 (29) |
| **Due to COVID-19** | 57 (29) |
| Novel coronavirus/COVID-19 has severely damaged the economy | 44 (77) |
| Slowing the spread of the pandemic is more important than damage to the economy | 38 (67) |
| Public health should be prioritized over opening the economy | 38 (67) |
| **Not Due to COVID-19** | 137 (71) |
| Novel coronavirus/COVID-19 has severely damaged the economy | 109 (80) |
| Slowing the spread of the pandemic is more important than damage to the economy | 82 (60) |
| Public health should be prioritized over opening the economy | 84 (61) |
| **Retired** | 164 (24) |
| Novel coronavirus/COVID-19 has severely damaged the economy | 144 (88) |
| Slowing the spread of the pandemic is more important than damage to the economy | 112 (68) |
| Public health should be prioritized over opening the economy | 122 (74) |

**S Table 4:** Participants Confidence in Healthcare Providers and Healthcare and Government Officials

| How much confidence do you have in each of these organizations? | Feb. 2020<br>Survey<br>No. (%) | May 2020<br>Survey<br>No. (%) | Chi-square p-value |
| --- | --- | --- | --- |
| Your own doctor | 534 (76) | 471 (73) | 0.24 |
| Local/County health department | 467 (67) | 411 (64) | 0.28 |
| State health department | 466 (67) | 419 (65) | 0.47 |
| Centers for Disease Control and Prevention (CDC) | 568 (80) | 430 (66) | **<0.01** |
| Food and Drug Administration (FDA) | 416 (60) | 351 (55) | 0.05 |
| White House | 211 (30) | 228 (35) | 0.07 |
| Department of Health and Human Services | 427 (62) | 313 (49) | **<0.01** |
| National Institute of Health | 496 (73) | 357 (57) | **<0.01** |
| Professional Organizations (eg: AMA or IDSA) | 509 (74) | 390 (61) | **<0.01** |

**S Table 5:** Risk Perception Score items with scoring guide (May 2020)

| **No.** | **Item** | **Score** |
| --- | --- | --- |
| 1 | My health will be severely damaged if I contract novel coronavirus |  |
|  | • Yes | 1 |
|  | • No | 0 |
| 2 | I think novel coronavirus is more severe than flu |  |
|  | • Yes | 1 |
|  | • No | 0 |
| 3 | Even if I fall ill with another disease, I will not go to the hospital because I will get novel coronavirus |  |
|  | • Yes | 1 |
|  | • No | 0 |
| 4 | Novel coronavirus will inflict serious damage on my community |  |
|  | • Yes | 1 |
|  | • No | 0 |
| 5 | Novel coronavirus will spread widely in the United States |  |
|  | • Yes | 1 |
|  | • No | 0 |
| 6 | I am more likely to get the novel coronavirus than other people |  |
|  | • Yes | 1 |
|  | • No | 0 |
| 7 | I believe I can protect myself against the novel coronavirus |  |
|  | • Yes | 0 |
|  | • No | 1 |
| 8 | I believe I can protect myself against the novel coronavirus better than other people |  |
|  | • Yes | 0 |
|  | • No | 1 |
| 9 | I am very concerned about this pandemic |  |
|  | • Yes | 1 |
|  | • No | 0 |
| 10 | I expect this pandemic to get larger |  |
|  | • Yes | 1 |
|  | • No | 0 |

**S Table 6:**
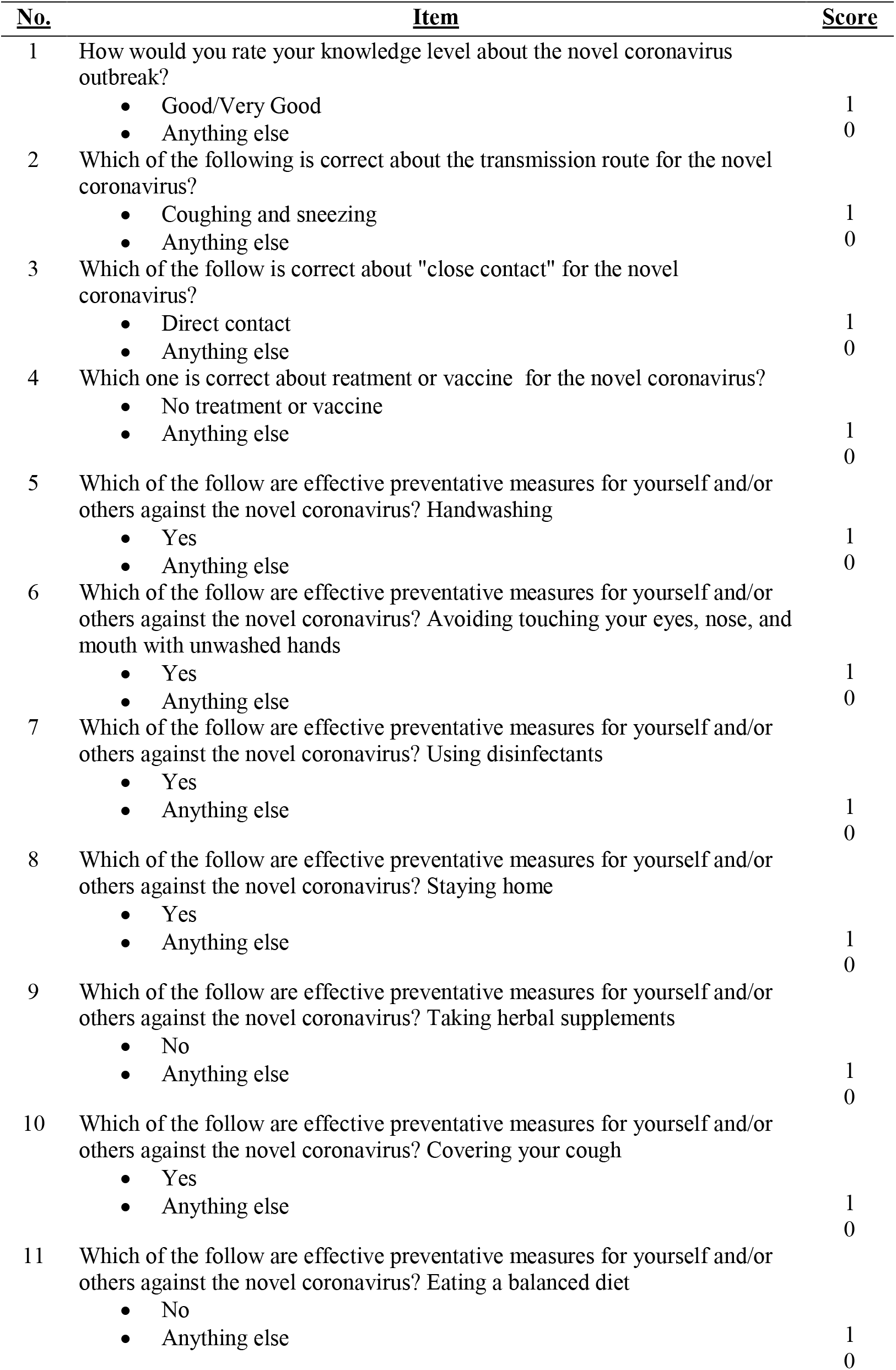
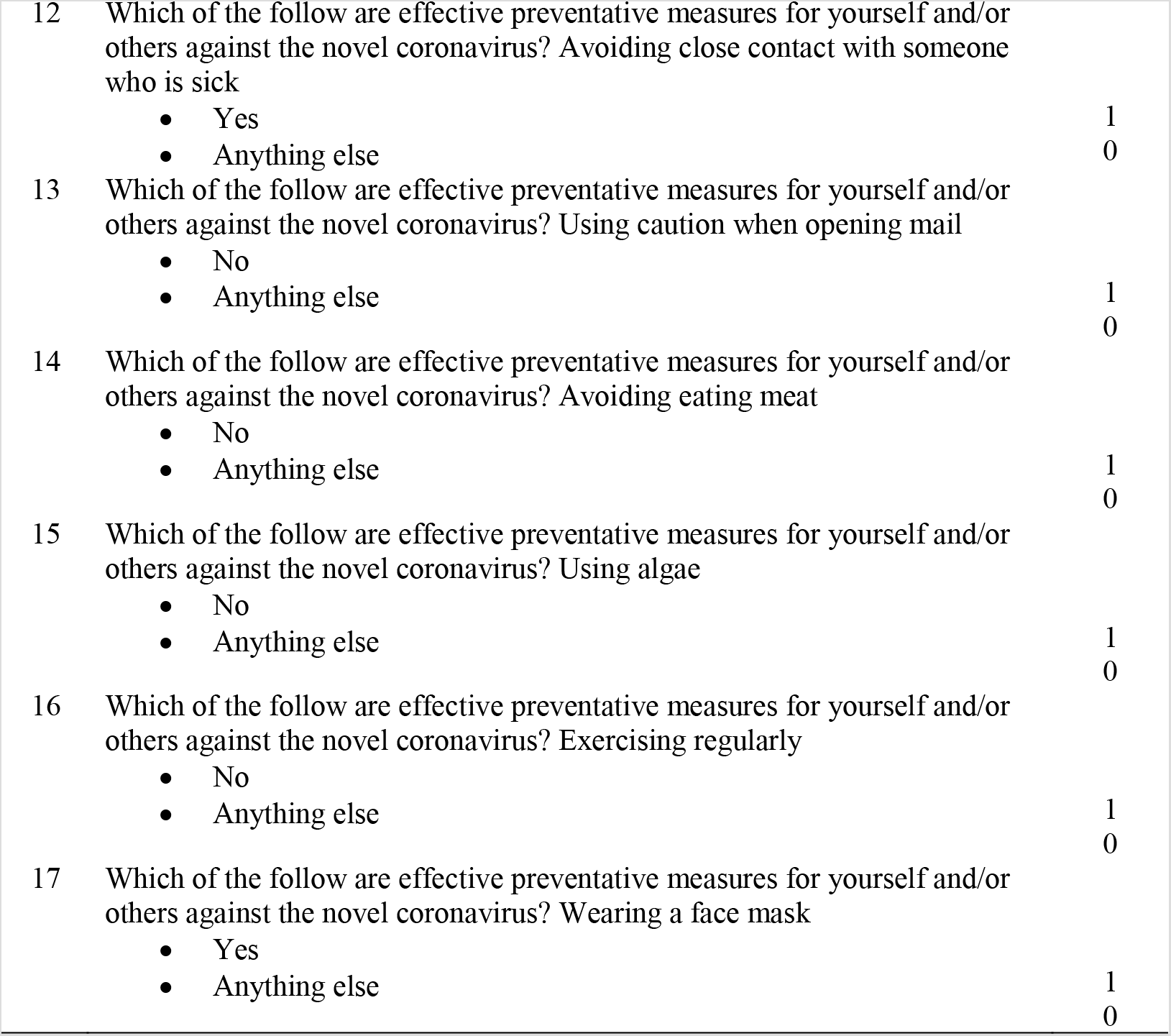
Risk Perception Score items with scoring guide (May 2020)

**S Figure 1:**
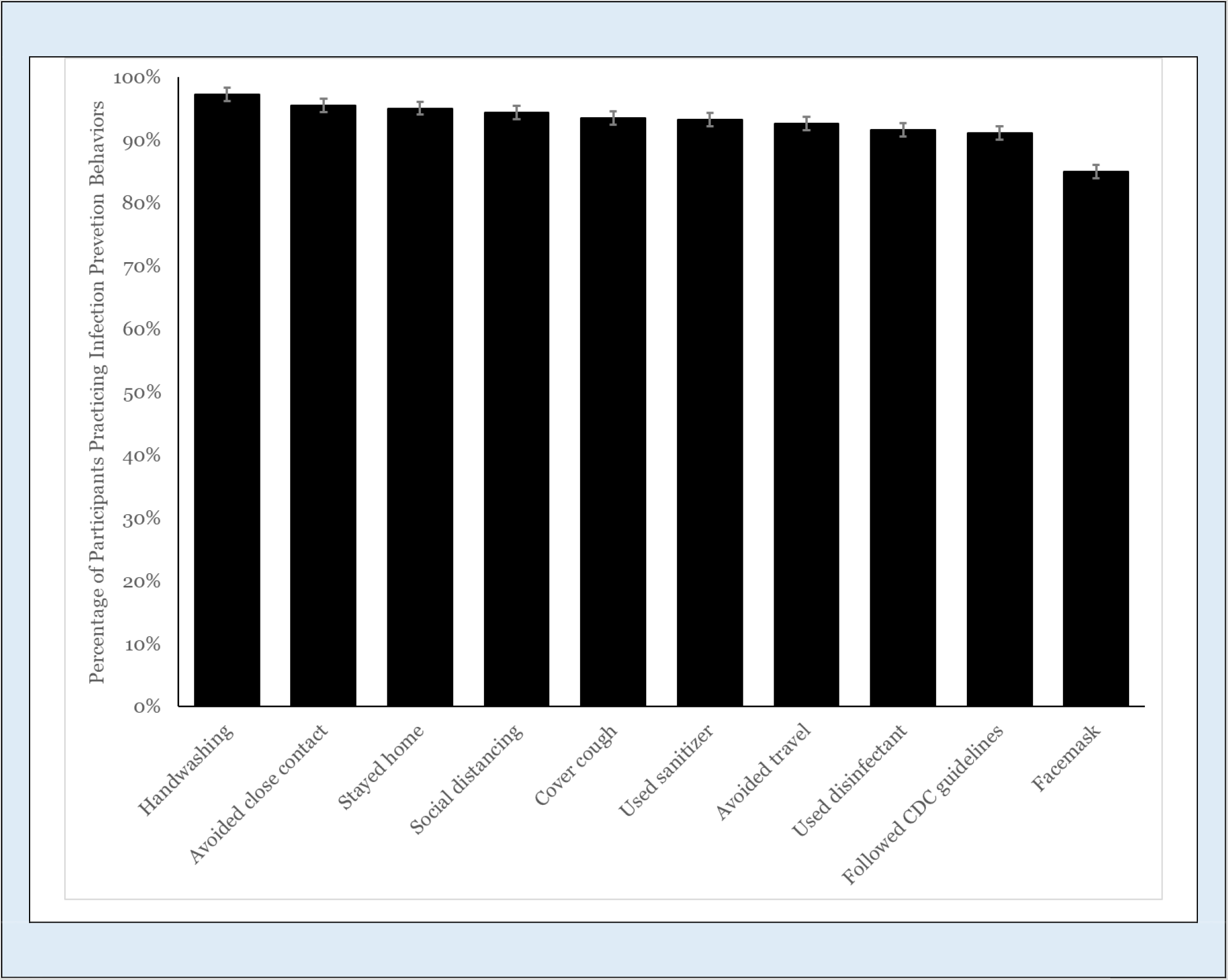
Percent of sample who participated in CDC recommended infection prevention strategies

**S Figure 2:**
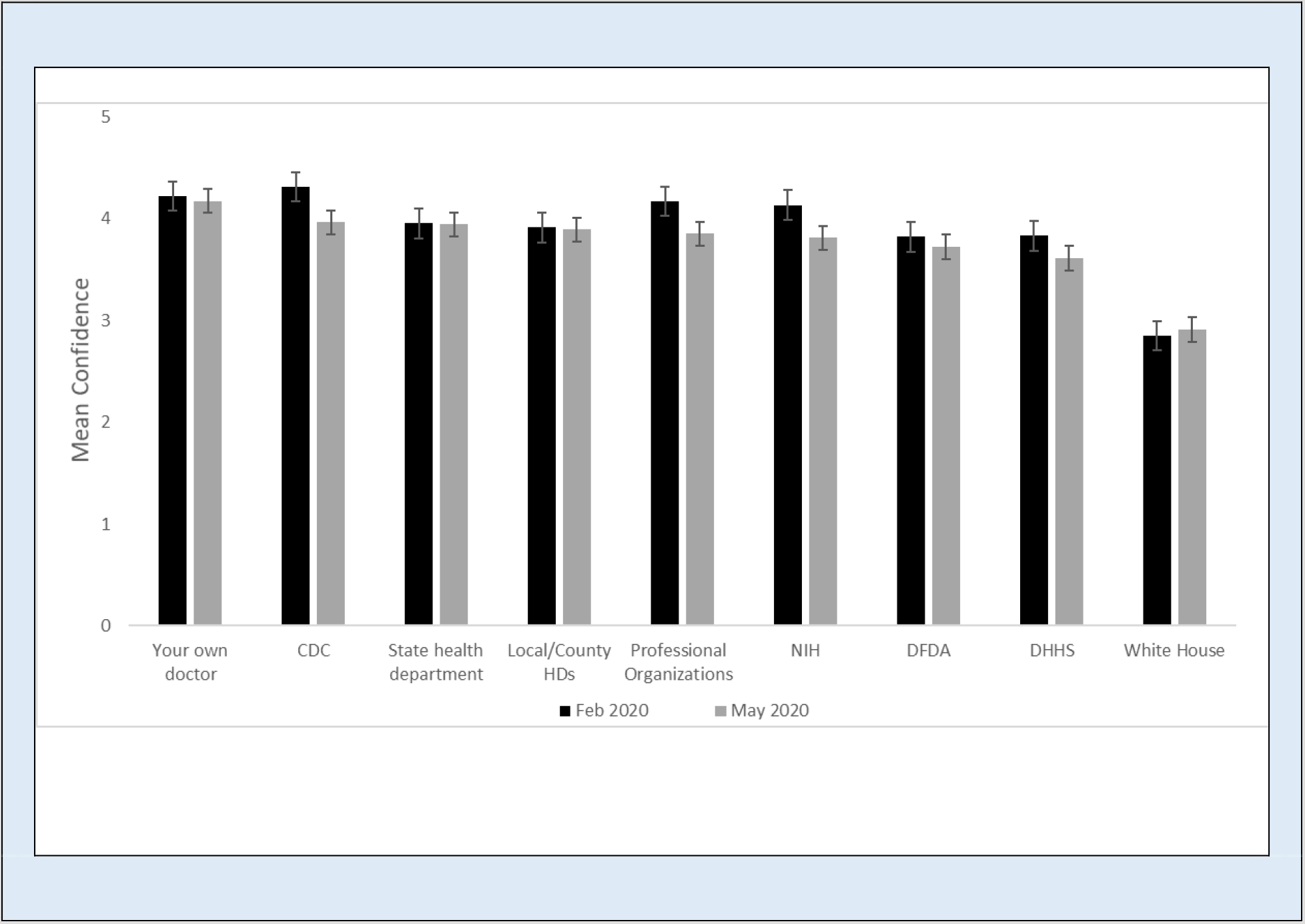
Participants’ mean confidence in each organization comparing survey result from February to May 2020

**S Figure 3:**
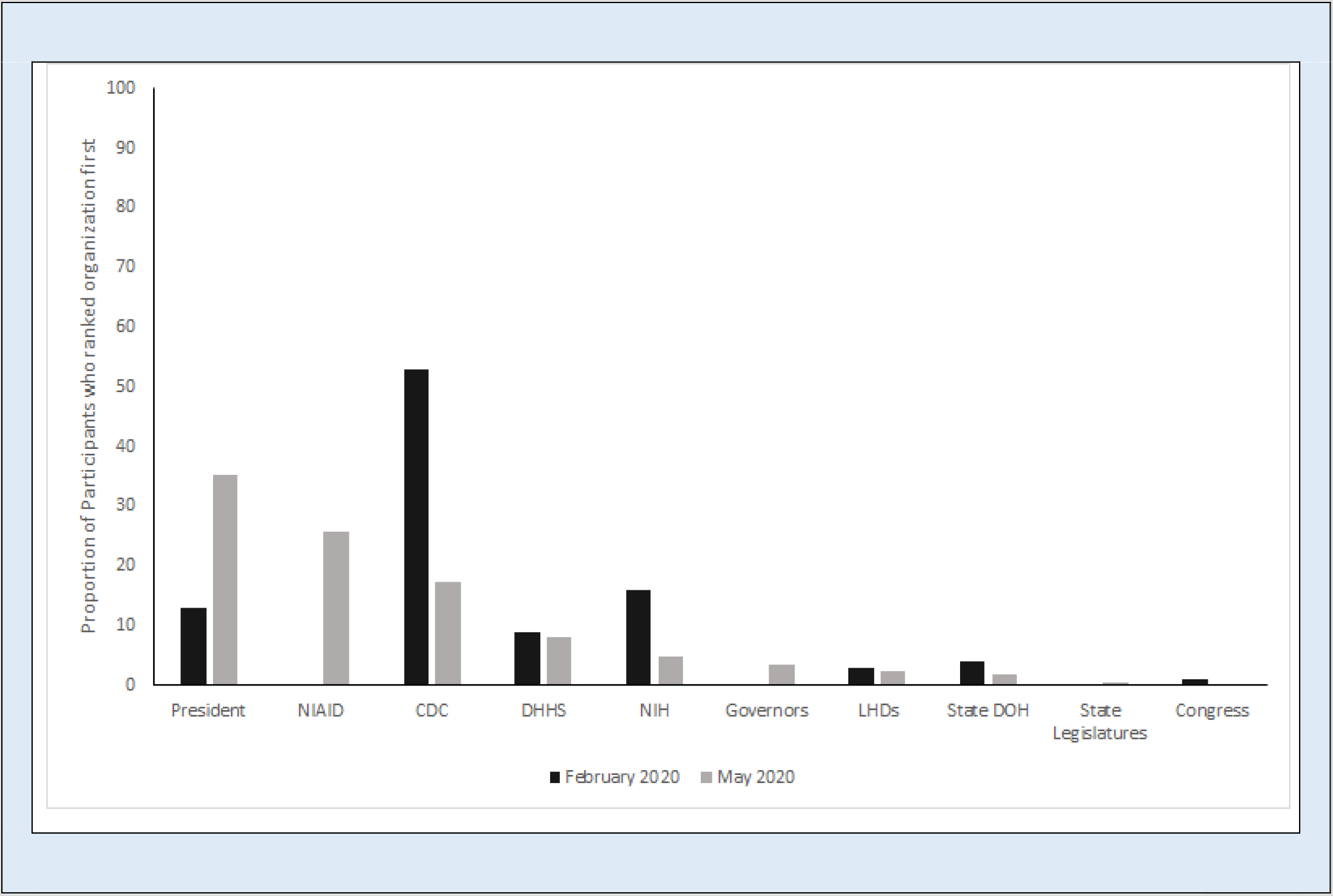
Comparison from February to May of who participants thought should lead the U.S. response to the COVID-19 pandemic

## Notes

### Competing Interest Statement

The authors have declared no competing interest.

### Author Declarations

Yale University Institutional Review Board approved this study (IRB protocol number: 2000027891).

